# The role of sub-genomic RNA in discordant results from RT-PCR tests for COVID-19

**DOI:** 10.1101/2021.12.14.21267750

**Authors:** Noah B. Toppings, Lisa K. Oberding, Yi-Chan Lin, David Evans, Dylan R. Pillai

**Affiliations:** Department of Microbiology, Immunology, and Infectious Diseases, University of Calgary, Calgary, AB, Canada; Department Pathology and Laboratory Medicine, University of Calgary, Calgary, AB, Canada; Department of Medical Microbiology & Immunology and Li Ka Shing Institute of Virology, University of Alberta, Edmonton, Alberta, Canada; Clinical Section of Microbiology, Alberta Precision Laboratories, Calgary, AB, Canada; Clinical Section of Infectious Diseases, Department of Medicine, University of Calgary, Calgary, AB, Canada

## Abstract

Reverse transcription-PCR (RT-PCR) is the standard method of diagnosing COVID-19. An inconclusive test result occurs when one RT-PCR target is positive for SARS-CoV-2 and one RT-PCR target is negative within the same sample. An inconclusive result generally requires retesting. One reason why a sample may yield an inconclusive result is that one target is at a higher concentration than another target. It was hypothesized that concentration differences across targets may be due to the transcription of sub-genomic RNA, as this would result in an increase in the concentration of gene targets near the 3’ end of the SARS-CoV-2 genome. A panel of six digital droplet (dd)PCR assays was designed to quantitate the ORF1, E-gene, and N-gene of SARS-CoV-2. This panel was used to quantify viral cultures of SARS-CoV-2 that were harvested during the eclipse phase and at peak infectivity in such a way as to maximize gene-to-gene copy ratios. Eleven clinical nasopharyngeal swabs were also tested with this panel. In culture, infected cells showed higher N-gene/ORF1 copy ratios than culture supernatants. Both the highest specific infectivity (copies/pfu) and the highest differences between gene targets were observed at 6 hours post-infection (eclipse phase) in infected cells. The same trends in the relative abundance of copies across different targets observed in infected cells was observed in clinical samples, though trends were more pronounced in infected cells. This study showed that a greater copy number of N-gene relative to E-gene and ORF1 transcripts could potentially explain inconclusive results for some RT-PCR tests on low viral load samples. The use of N-gene RT-PCR target(s) as opposed to ORF1 targets for routine testing is supported by this data.

**Author Summary:** This paper provides insight into a drawback of the standard method of testing for COVID-19 (RT-PCR). The results presented here propose an explanation for why inconclusive results sometimes occur with this method. These results can aid microbiologists in the interpretation of inconclusive test results. These results can also aid in decisions about which COVID-19 test a laboratory should use, as there are a plethora of options available. This is important because this standard testing method will remain a critical tool – globally – for managing the COVID-19 pandemic and any future viral pandemics and epidemics. Thus, it is important to investigate every facet of the testing method. The findings presented here are applicable to any virus which makes sub-genomic transcripts as part of its life cycle. Trends observed in viral cultures are presented alongside the same trends observed in clinical samples. Unlike similar papers in the field, this paper did not strive to develop a new methodology or tool.

## Introduction

RT-PCR is the standard method of diagnosing COVID-19 in a laboratory (1–4). RT-PCR tests for COVID-19 qualitatively detect SARS-CoV-2 RNA from patient samples (1–4). SARS-CoV-2 is a +sense RNA virus of the *Coronaviridae* family with a genome approximately 29,903 nucleotides long (5,6). Diagnostic RT-PCR tests which detect SARS-CoV-2 RNA usually identify two or three individual RT-PCR targets in order to increase one’s confidence in the results, control for possible errors, and provide redundancy in the case of mutations which effect primer binding (7–10). Thus, the qualitative result of the RT-PCR test is reached by interpreting the results of all the individual RT-PCR targets. Table 1 is an example of an interpretation guide for a typical RT-PCR test for COVID-19.

**Table 1.** A hypothetical example of a COVID-19 RT-PCR results interpretation guide. A control assay which targets a human gene, or externally added control material, is often run to ensure that sample collection and nucleic acid extraction was performed properly and to ensure that amplification is not being inhibited by the sample.

| **RT-PCR**<br>**Target** | **Assay Result** |  |  |  |  |
| --- | --- | --- | --- | --- | --- |
| E gene | - | + | - | + | - |
| RdRP | - | + | - | - | + |
| RNase P<br>(internal<br>control) | + | +/- | - | +/- | +/- |
| COVID-19<br>Test Result | COVID-19<br>Not<br>Detected | COVID-19<br>Positive | Invalid | Inconclusive | Inconclusive |

For each clinical sample, all the SARS-CoV-2 RT-PCR targets usually yield the same qualitative result and yield similar Ct values. This is because viral replication produces full-length SARS-CoV-2 genomes which contain all genes in equal proportions (Figure 1)(11,12). However, sometimes only a single SARS-CoV-2 assay will yield a positive result, making the test result inconclusive. Samples with low viral loads are more likely to yield an inconclusive result in this way (10). The biological and clinical significance of low viral load (high Ct value) samples is under investigation as some reports suggest these samples are rarely infectious (13,14).

**Figure 1.**
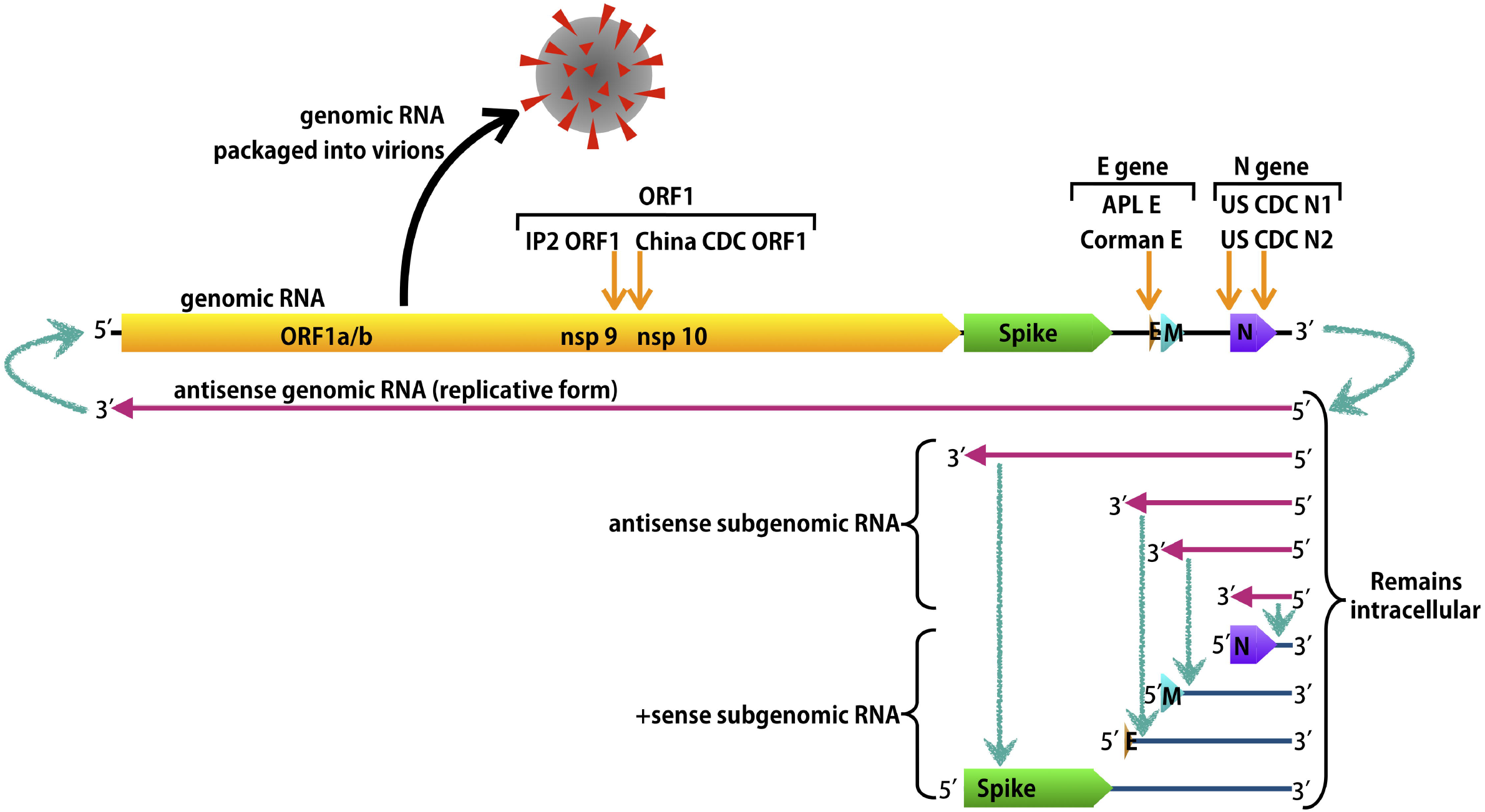
Simplified drawing of SARS-CoV-2 genome replication and transcription showing a subset of the canonical sub-genomic RNAs. The approximate locations of the ddPCR assays used in this study are indicated. After infecting a cell, the SARS-CoV-2 genome gets transcribed to make either (full length) antisense genomic RNA (the replicative form) or antisense sub-genomic RNA (11,12). Sub-genomic RNAs are produced via discontinuous transcription and are considered to be ‘nested’ because they all start from the same 3’ end of the genome (11,12). Antisense genomes can then be transcribed to make +sense genomes which can complete this cycle again or be encapsidated inside a virion (11,12,32). Antisense sub-genomic RNA can be transcribed to make +sense sub-genomic RNA which can be translated to make the protein encoded at the 5’ end of the sub-genomic RNA molecule (11,12). Diagnostic primer sets for RT- and ddPCR amplify all RNA species containing their target sequence, regardless of whether the RNA is sense or antisense, genomic or sub-genomic.

Some studies have reported the prevalence of inconclusive COVID-19 RT-PCR test results. Vogels *et al*. observed that 7.9% of samples positive for at least one RT-PCR target were inconclusive (not positive for both targets) (15). Out of 286 samples tested, Jawade *et al*. observed 19 (6.64%) inconclusive samples and 126 (44.1%) positives (16). With the specific kit Jawade *et al*. used, a sample was inconclusive only when the RdRP target was negative and the E gene target was positive (16,17).

When a sample is deemed to be inconclusive, retesting of the sample is generally recommended (2,17). Retesting consumes valuable time and reagents; decreasing testing throughput. Thus, it is advantageous to understand the causes of inconclusive test results so they can be minimized.

There are various reasons why an RT-PCR test might yield an inconclusive result. Reasons include, differences in sensitivity between primer/probe sets, mistakes setting up the reaction, primer-template mismatches caused by a mutation in the virus, differences in the template concentrations of different targets, among other factors, and a combination of these factors.

As an example, Vogels *et al*. determined that the US CDC N1 primer/probe set was more sensitive than the US CDC N2 primer/probe set (15). Using a Ct value positivity cutoff of 38, Vogels *et al*. tested 172 clinical samples with the US CDC N1 and N2 primer/probe sets (15). From this set, 58/172 (33.7%) were positive while 5/172 (2.9%) were inconclusive, representing 7.9% of samples positive for at least one target (15). All 5 inconclusive samples were positive for the more sensitive N1 target and negative for the N2 target (15).

Documented examples of specific SARS-CoV-2 mutations causing inconclusive RT-PCR test results include the C26340U transition (8) and the Δ69-70 deletion in the spike protein of the B.1.1.7 lineage (18). These mutations caused failure of the E gene target with the Roche cobas test (8), and the spike gene target using the ThermoFisher TaqPath probe (18), respectively. This study focuses solely on inconclusive results caused by differences in the template concentrations of different targets.

We hypothesized that discrepancies in the qualitative result of different RT-PCR assays on SARS-CoV-2-suspected clinical samples can largely be explained by a greater abundance of gene copies near the 3’ end of the genome due to the transcription of sub-genomic RNA. To test this hypothesis, a panel of six ddPCR assays was designed to quantify the abundance of different SARS-CoV-2 RT-PCR targets. Next, SARS-CoV-2 was cultured *in vitro* to observe the relative abundance of different targets under ideal conditions. Finally, a set of clinical samples was tested to observe trends in the relative abundance of different targets. This study showed that greater copies of the N-gene relative to the E gene and ORF1 could potentially explain inconclusive results of some RT-PCR assays on low viral load samples.

## Results

### Droplet Digital PCR Methodology Validation

To quantify different regions of the SARS-CoV-2 genome, six pre-existing RT-PCR assays intended for routine COVID-19 diagnostic testing were adapted to ddPCR. In the case of the US CDC N1 and N2 targets, these had already been adapted by Bio-Rad (19). As the objective of this study was to better understand routine diagnostic RT-PCR testing, no primer sets for research purposes were used or designed.

Differences in the sensitivity of various primer/probe sets could give the false appearance of a difference in target gene abundance. However, this was ruled out by using the Exact Diagnostics SARS-CoV-2 (quantitative) Standard. The counts using this standard for each target are shown in Figure S2. Assays that did not quantitate the standard accurately were considered non-quantitative and were excluded from this study. This experiment used two ddPCR assays targeting each of the following open-reading frames: nucleocapsid (N), envelope (E), and ORF1ab. Differences in the relative abundance of different SARS-CoV-2 targets (as determined by ddPCR counts) could be the result of unequal transcription of targets (sub-genomic RNA), or the unequal degradation of RNA targets.

As this study used ddPCR to investigate the performance of RT-PCR, it was important to evaluate the performance of ddPCR relative to RT-PCR. To this end, RT-PCR and ddPCR was performed on three replicate serial 10-fold dilutions of a clinical sample (Figure 2). Six standard curves were prepared from this data (Figure 3). These curves all showed highly linear relationships between RT-PCR Ct values and log_10_(copies/µL) from ddPCR across the dilution series with only one adjusted R^2^ value being below 0.97 (Figure 3). The exception being the IP2 ORF1 target due to high variance from the RT-PCR (Figures 2 and 3). The IP2 ORF1 RT-PCR coefficients of variation were on average 6.84-fold higher than for the other five RT-PCR targets. However, all six linear regressions had overlapping 95% CI’s for both the coefficient and the constant (Figure 3). This suggests that these two techniques yield highly concordant results and supports applying conclusions from ddPCR to RT-PCR.

**Figure 2.**
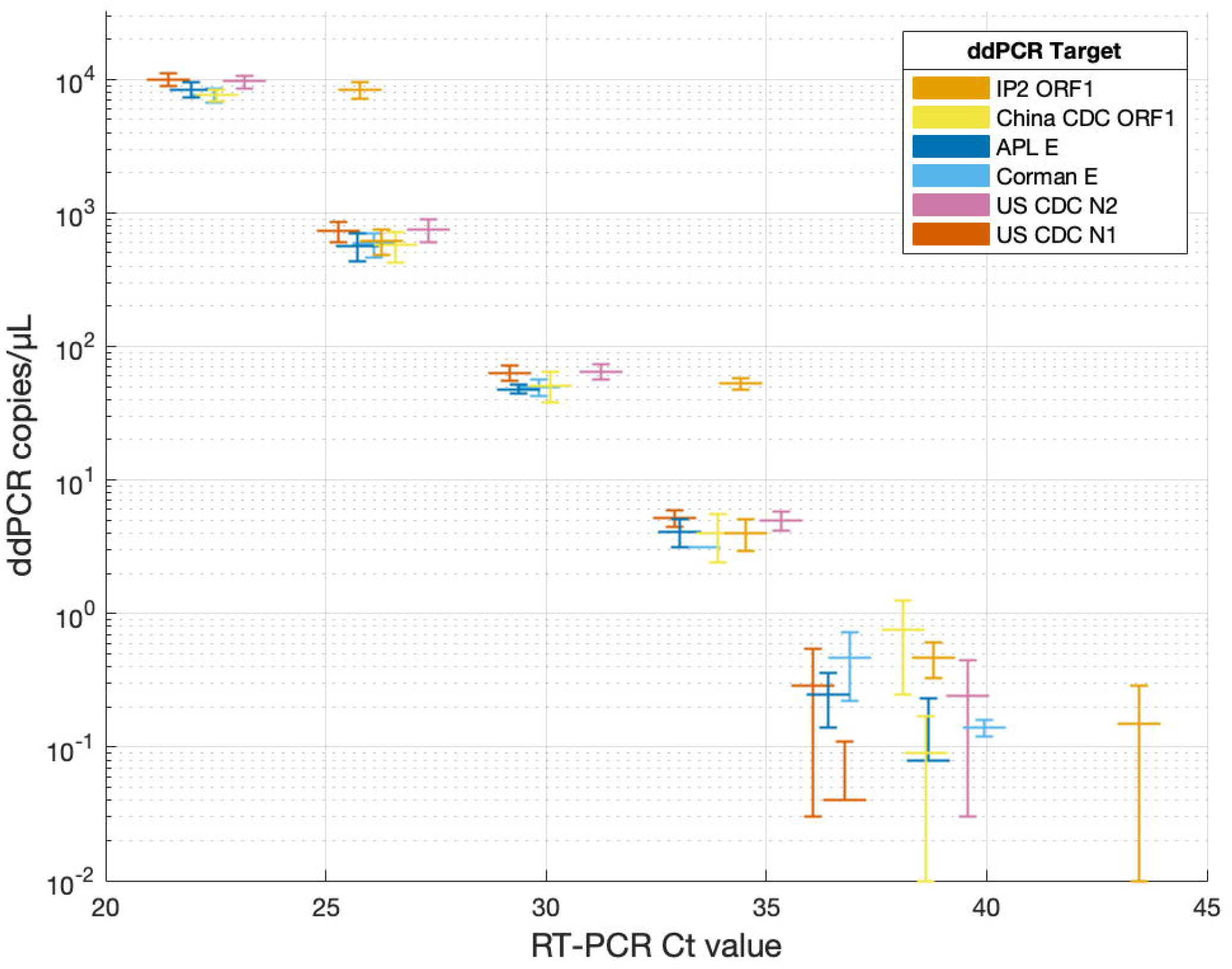
Triplicate serial 10-fold dilution of a clinical nasopharyngeal swab (sample 1) tested by RT-PCR and quantified by ddPCR. Dilutions from 10-fold to 1 million-fold are shown. Error bars show ± 1 standard deviation from the arithmetic mean of triplicate ddPCR counts. Triplicate RT-PCR Ct values were averaged to simplify interpretation. RT-PCR reactions which did not amplify (only at the 1 million-fold dilution) were excluded from the analysis. The 1 million-fold dilution for the US CDC N2 target is not shown because the RT-PCR did not amplify for any replicates.

**Figure 3.**
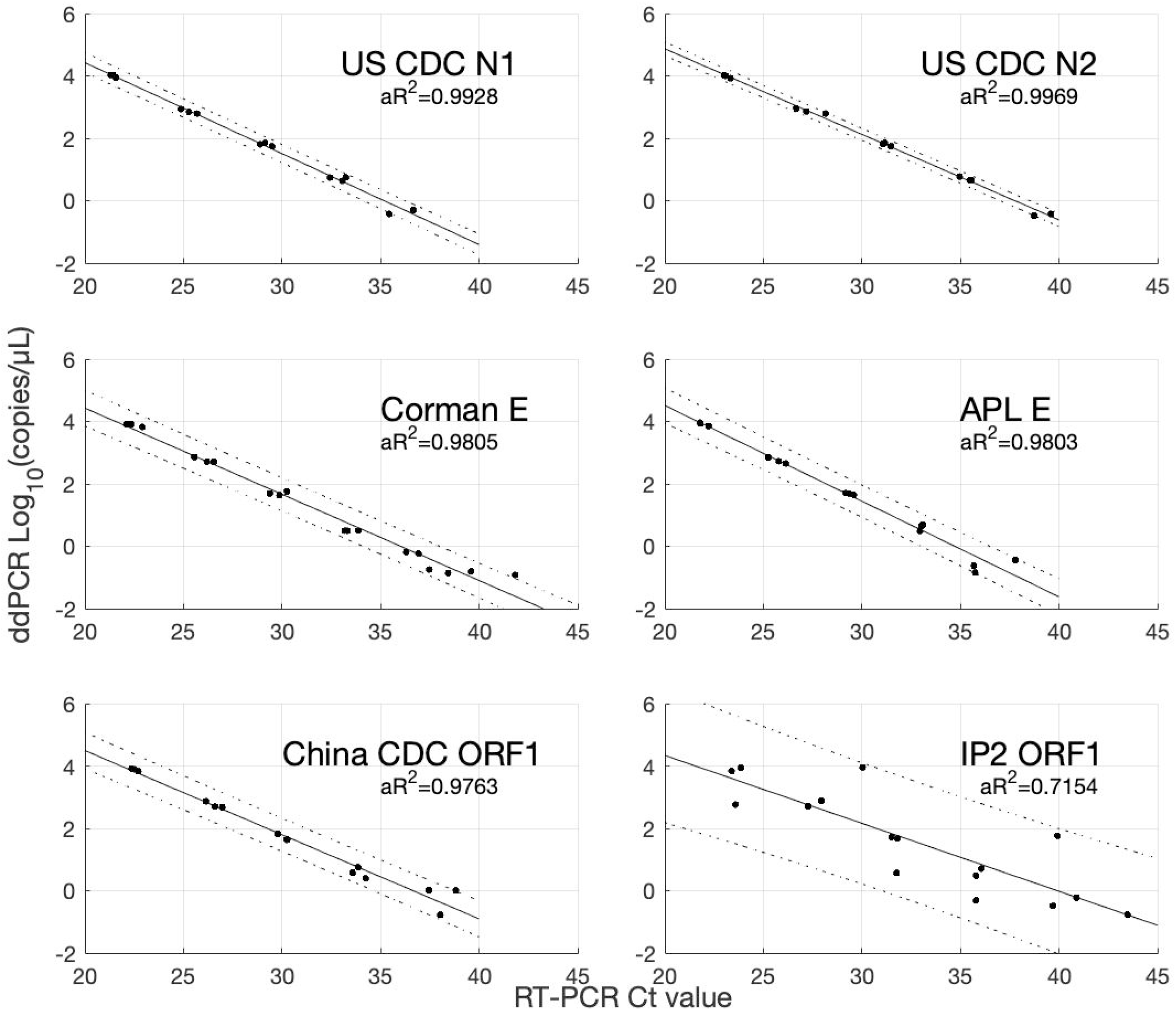
Triplicate serial 10-fold dilution of a clinical nasopharyngeal swab tested by RT-PCR and quantified by ddPCR. Linear regression with 95% prediction bands and adjusted R^2^ shown. Dilutions from 10-fold to 1 million-fold are shown. If either the RT-PCR or the ddPCR reaction did not amplify, the replicate was excluded from the analysis.

The RNase P gene of Vero CCL81 cells differs from that of human RNase P such that there are four primer-template mismatches; two in each primer (20–24). This was predicted to greatly decrease the sensitivity of this primer/probe set (25). Thus, RNase P ddPCR counts from Vero CCL81 cells were not used for quantitative comparisons.

### SARS-CoV-2 *in vitro* Cultures

Vero CCL81 cells were infected with SARS-CoV-2 at a multiplicity of infection of 0.01 and grown for 48 hours. A sample of the culture supernatant and the adherent cells was taken at 6 and 48 hours post-infection (HPI). The 6 HPI time point occurs just after the eclipse phase of the virus (26) while the 48 HPI time point occurs at peak infectivity (our unpublished data). The cell lysate was expected to be the exclusive site of viral replication while the culture supernatant was expected to contain released virions and minimal cell contents (27). Figure 4A shows the infectivity of viral cultures over time, with overlapping infectivity between supernatants and cell lysates at 6 HPI and around a 4log_10_ increase in infectivity by 48 HPI with cell lysates more infectious than the supernatants at 48 HPI. Specific infectivity (copies/pfu) was highest in cell lysates and at 6 HPI opposed to 48 HPI (Figure 4B).

**Figure 4.**
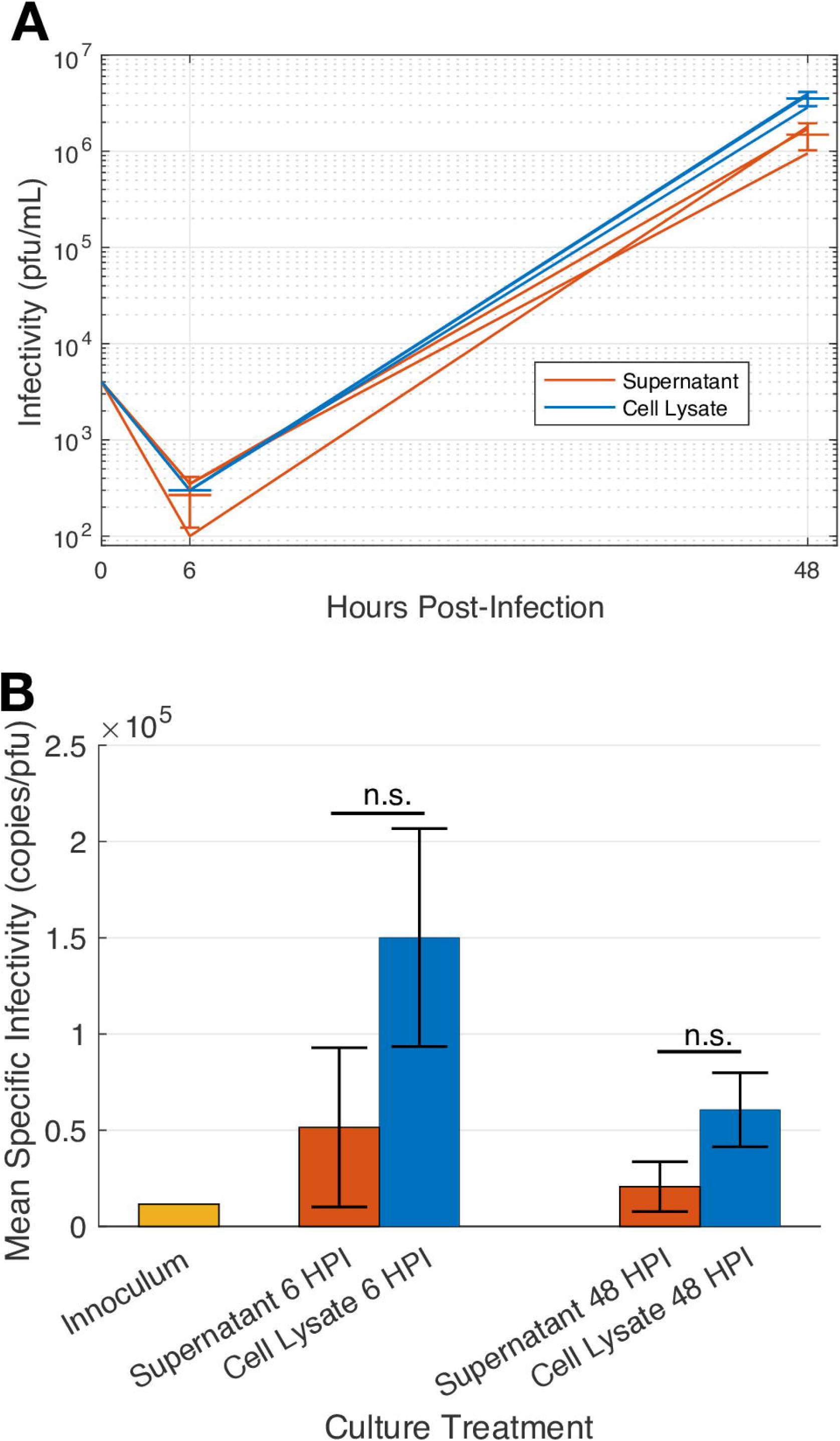
**A) Growth curve of SARS-CoV-2 cultured in Vero CCL81 cells over 48 hours**. The infectivity of the hour zero time point is calculated based on the intended number of pfu in the inoculum at the start of the infection (10,000 pfu/ 2.5 mL media) (multiplicity of infection = 0.01). **B) Mean specific infectivity of SARS-CoV-2 cultures is highest in cell lysates and just after the eclipse phase of growth**. The copies/mL of all six ddPCR targets was averaged and then divided by pfu/mL to yield mean specific infectivity. Supernatants and cell lysates were compared with the Mann-Whitney U-test (n.s.: p>0.05). HPI: hours post-infection. Error bars show ± 1 standard deviation from the arithmetic mean of 3 replicates. The inoculum was not replicated (n=1).

Similar trends observed for specific infectivity were also observed in the relative abundance of different gene targets (Figure 5). The greatest differences in copies between targets was observed in cell lysates, opposed to supernatants (Figure 5). Between the two supernatants, the greatest differences between targets were observed at 6 HPI while for cell lysates, however, the greatest differences were observed at 48 HPI (Figure 5). Within a single target, higher variances between replicates was observed at 6 HPI rather than at 48 HPI (Figure S3).

**Figure 5.**
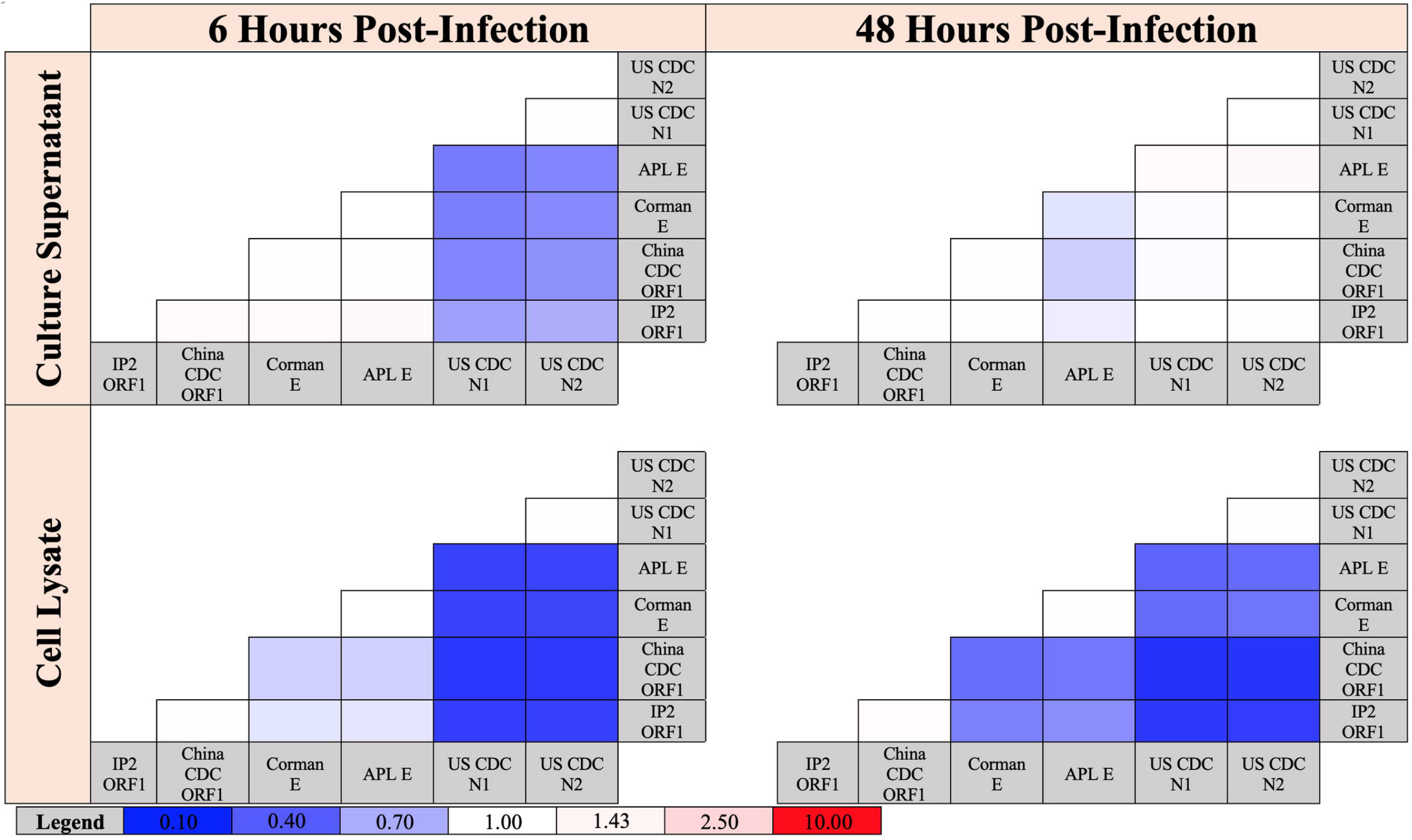
Relative abundance of each ddPCR target for SARS-CoV-2 cultures at each time point, and for each culture fraction. For each sample, the copies of each target, indicated in the left column, was divided by the copies of each target, indicated in the bottom row. Each cell was colored based on the arithmetic mean of 3 replicates.

In both the supernatant and cell lysate at 6 HPI, the highest counts were from the N gene targets with relatively similar counts from the E gene and ORF1 targets (Figure 5). In the supernatant at 48 HPI, the APL E gene target yielded the highest counts with relatively similar counts between the remaining 5 targets. In the cell lysate at 48 HPI, the highest and lowest counts were from the N-gene and ORF1 targets, respectively, with the two E-gene targets yielding intermediate counts (Figure 5). In summary, this viral culture experiment shows that under various conditions (especially in infected cells or during the eclipse phase) different SARS-CoV-2 gene targets are present at different concentrations in a manner consistent with the transcription of sub-genomic RNA. The question then became whether or not these same trends observed *in vitro* could be observed *in vivo* in clinical samples from people infected with SARS-CoV-2.

### Analysis of Clinical Nasopharyngeal Swabs

To assess whether the trends observed *in vitro* could also be observed *in vivo*, eleven nasopharyngeal swabs from people confirmed to have COVID-19 were tested with the panel of six ddPCR targets. The range of RT-PCR Ct values of the eleven clinical nasopharyngeal swabs was 19.04 to 31.95 (US CDC N2 target) (Table S1). The absolute abundance, and relative abundance, of each ddPCR target for the eleven clinical samples tested is shown in Figures 6 and 7, respectively. Clinical samples yielded the highest counts with the N-gene targets (Figure 7). The E-gene targets and IP2 ORF1 yielded similar counts while the China CDC ORF1 count was the lowest across the samples (Figure 7). Thus, the same trends in the relative abundance of different ddPCR targets observed in viral cultures was also observed in clinical samples.

**Figure 6.**
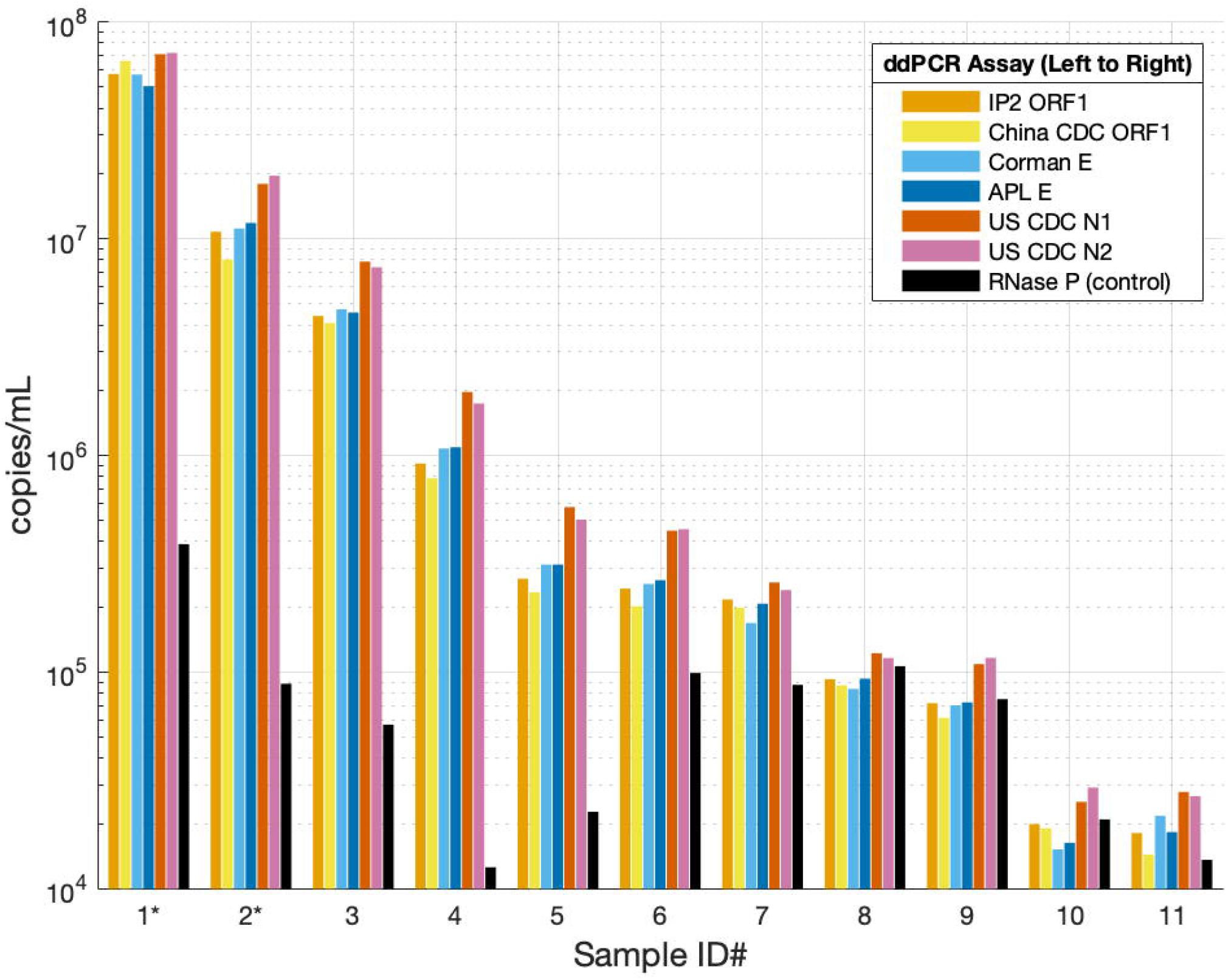
Clinical nasopharyngeal swabs quantified by ddPCR. *The counts shown for samples 1 and 2 are from a 1000-fold dilution (corrected for the dilution factor) due to signal overload when quantifying the undiluted sample.

**Figure 7.**
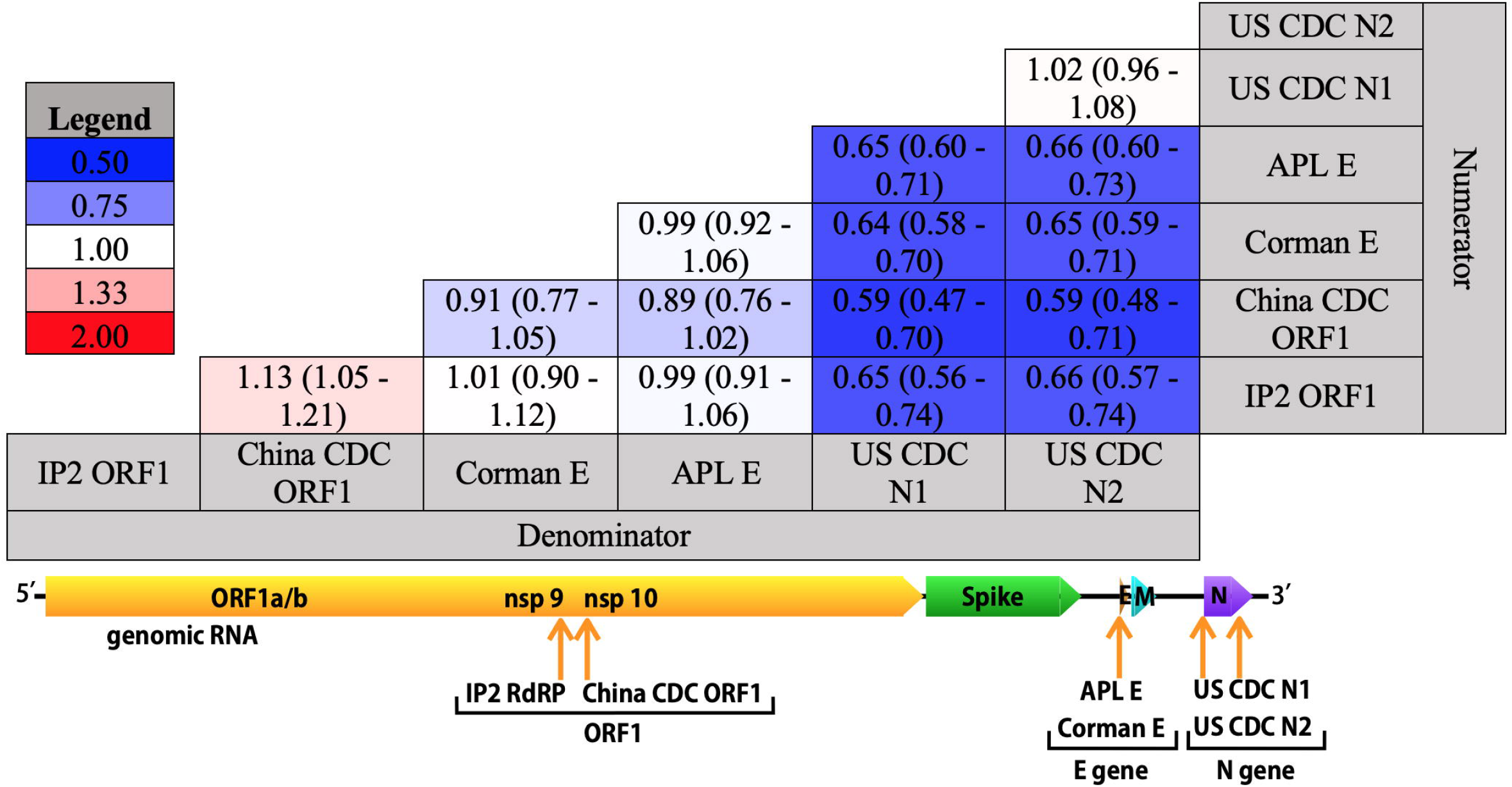
Relative abundance of each ddPCR target in clinical samples. Mean (95% CI) copy ratios (n = 11) between all SARS-CoV-2 ddPCR assays for each clinical nasopharyngeal swab. For each sample, the copies of each target, indicated in the left column, was divided by the copies of each target, indicated in the bottom row. Following this, the arithmetic mean and 95% CI of the eleven ratios were calculated for each cell and a shade of blue or red was assigned based on the value.

For a detailed comparison of the relative abundance of ddPCR targets in clinical samples to that of cultures, the mean copy ratios from the cultured samples were subtracted from the mean copy ratios from the clinical sample set (Figure 8). The more similar the copy ratios were, the paler the matrix (Figure 8). The N-gene/ORF1 ratios were higher in the cell lysates than in clinical samples (Figure 8). The same held true for the E-gene/ORF1 ratios (Figure 8). Visually, and numerically, the clinical samples were most similar to the culture supernatants at 6 HPI (eclipse phase) (Figure 8). The N-gene/ORF1 ratios in the culture supernatants at 48 HPI were less than the ratios in clinical samples (Figure 8). In summary, clinical samples had differences between target copies intermediate that of cell lysates and culture supernatants (Figure 8).

**Figure 8.**
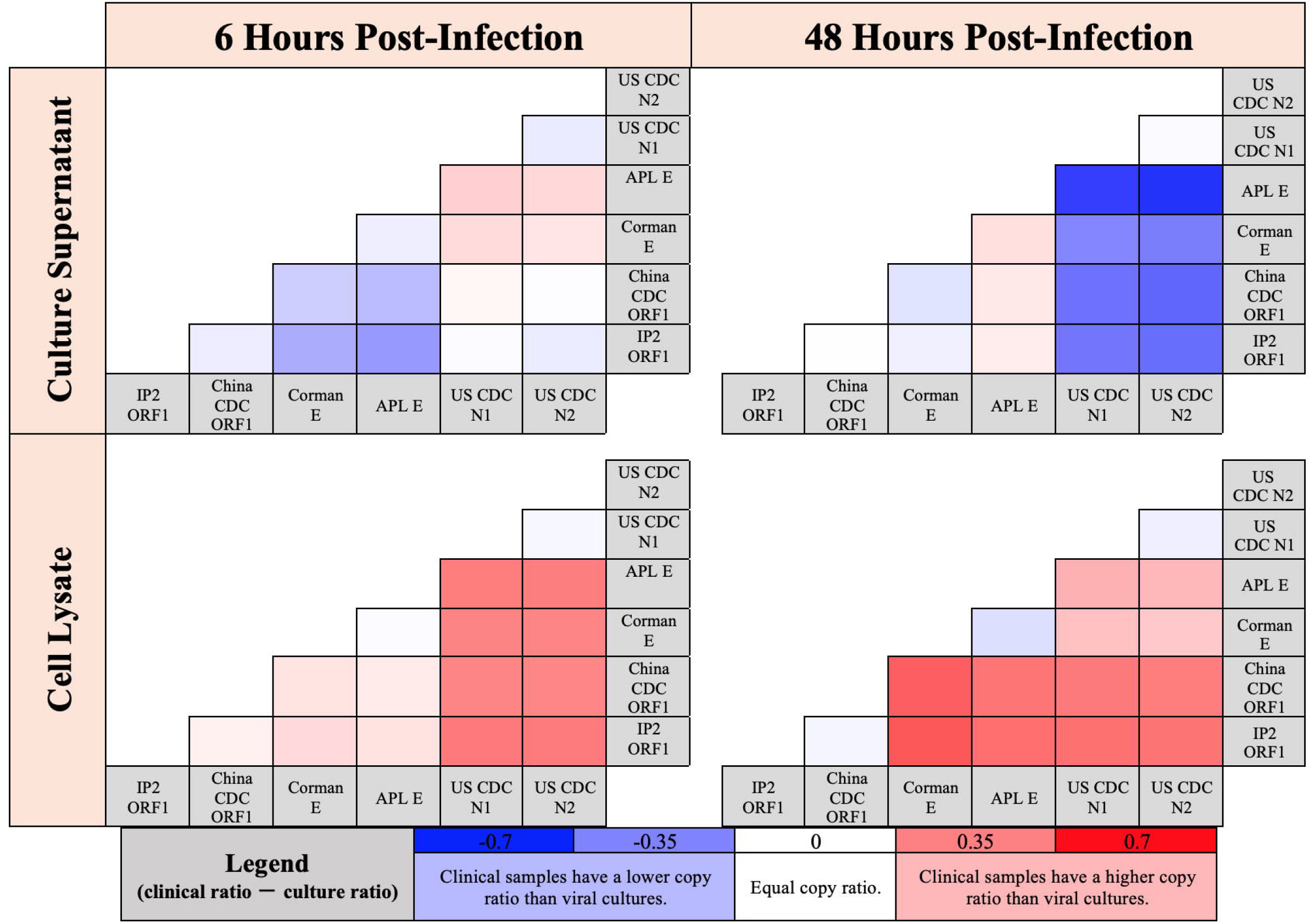
Comparison of clinical samples to viral cultures by the relative abundance of each ddPCR target. The mean copy ratios from the cultured samples (Figure 5) were subtracted from the mean copy ratios from the clinical sample set (Figure 7) (clinical – culture).

## Discussion

The results presented here propose an explanation for some inconclusive COVID-19 RT-PCR test results. Specifically, when inconclusive RT-PCR results occur due to an ORF1 target being negative and an N gene target being positive. In such cases, this may be due to fewer ORF1 copies in the sample. Data support the use of diagnostic RT-PCR assays targeting the N-gene exclusively, as opposed to targeting the E-gene or ORF1, for optimal sensitivity. This is because, on average, more N-gene copies were detected in clinical samples than ORF1 copies. However, the concentration of ORF1 copies in clinical and viral culture samples in this study was always above the limit of detection of ddPCR.

The reason why clinical samples may have more N-gene copies than ORF1 copies could be because clinical samples do not merely contain virions but contain infected cells which contain sub-genomic RNA encoding the N-gene. However, this study never specifically identified sub-genomic RNA because the diagnostic tests in question do not specifically identify sub-genomic RNA. Thus, it cannot be ruled out that unequal degradation of genomic RNA could explain the trends observed. All assays amplified their specific target regardless of whether it was in genomic, sub-genomic, sense, or antisense transcripts.

Similar studies have been published describing panels of optimized ddPCR assays for SARS-CoV-2 (28,29). However, the goal of this study was to understand a deficiency in RT-PCR tests, not to develop a tool for understanding SARS-CoV-2 biology nor was the goal to develop a diagnostic test for clinical use. Nonetheless, the trends of E-gene and N-gene copies identified here are congruent with the findings of Telwatte *et al*. (both papers) in that genes nearest the 3’ end of the genome have greater copy numbers than those in ORF1 (28,29). Consistent with the results presented here, Dimcheff *et al*. found – using RT-qPCR – that N-gene sub-genomic RNA was more detectable than E gene sub-genomic RNA after testing 185 clinical nasopharyngeal swabs (30). However, Dimcheff *et al*. reported a minimal difference between the mean Ct values of the US CDC N1 and Corman E RT-qPCR assays across all 185 samples (mean Ct of 25.6 S.D. 5.6 versus 25.76 S.D. 5.69, respectfully) (30). It is not clear if a sample-wise comparison of US CDC N1 and Corman E Ct values would show significantly lower Ct values for US CDC N1. Likewise, the mean copies/mL for US CDC N1 and Corman E across all clinical samples in this study were not close to significantly different (p = 0.84 with Mann-Whitney U test and p = 0.51 with two-sample t-test). Nonetheless, this study reported that US CDC N1 copies were significantly higher than Corman E copies on a sample-wise basis (Corman E/US CDC N1 = 0.65 95%C.I. 0.60-0.71) (Figure 7). Furthermore, both E-gene ddPCR targets had significantly lower counts than either N gene ddPCR target, with the E-gene/N-gene ratios ranging from 0.64 to 0.66 (Figure 7). This apparent disagreement could be partially resolved with a sample-wise analysis of Ct values or else it could be explained by the smaller sample size of this study or differences in the quantitative accuracy of ddPCR versus RT-PCR.

Marchio *et al*. performed a ddPCR study reminiscent of this study and obtained congruent results from clinical samples and cultured virus (31). In viral cultures at 24 HPI, the highest copy target was the N-gene followed by the E-gene and finally the IP2 ORF1 and IP4 ORF1 targets (31). Importantly, these results were observed across three different mammalian cell lines, none of which were used in this study (31). Thus, the results from viral culture presented here are clearly not restricted to a single cell line, increasing their relevance to understanding clinical samples (31). More N-gene positive droplets than ORF1 positive droplets in 9/16 clinical samples were observed, whereas this study observed higher N-gene counts compared to ORF1 counts in 11/11 clinical samples (Figure 6) (31). However, further analysis of this was not provided, preventing more comparisons between these two studies (31). Interestingly, ddPCR single-target positivity in five samples which were negative by RT-PCR was noted. Of these five, four were positive for only the N-gene target while one was only positive for the IP4 ORF1 target. Furthermore, the results of this study and Marchio *et al*. support the use of RT-PCR assays targeting the N-gene, opposed to targeting the E-gene or ORF1 (31). Hu *et al*. is the only study besides this study we have found to date which uses ddPCR to investigate inconclusive RT-PCR results (10). However, only two ddPCR targets were used in Hu *et al*. (10). These authors also conclude that N gene confers an advantage in terms of analytical sensitivity.

Alexandersen *et al*. used next-generation sequencing in combination with RT-PCR to understand the relative abundance of different SARS-CoV-2 sequences in clinical samples (32). From reads unambiguously mapped to subgenomic RNA transcripts, the transcript with the highest median read count across clinical samples was ORF7a, followed by the N-gene, then ORF8, and finally the E-gene (32). Again, this trend is consistent with the ddPCR results presented here. Diagnostic RT-PCR assays which specifically detect ORF7a were not identified in our review. Next-generation sequencing and subsequent bioinformatic pipelines have certain biases as these are longer workflows which often discard low quality or ambiguous sequences (33). By using the same diagnostic primer sets with ddPCR and RT-PCR, many of the biases introduced by next-generation sequencing and bioinformatic pipelines are controlled for in this study, thus increasing the relevance of these results to RT-PCR testing.

A limitation of this study is its small sample size of eleven clinical nasopharyngeal swabs. A larger number of clinical samples – and different types – should be tested in the future to confirm the conclusions made here. Furthermore, this study did not show that samples with less ORF1 copies than N gene copies would in fact be more likely to be ORF1 negative, N gene positive, in an RT-PCR test. Future studies could test for a relationship between copy number ratios and rates of inconclusive test results. A strength of this study is that it used two different ddPCR targets for each of the three ORFs of interest.

In conclusion, this study proposes a biological explanation for certain inconclusive RT-PCR results. Namely, sub-genomic RNA in clinical samples may increase the likelihood of an N gene target from being positive by RT-PCR. Further studies are required to validate this finding in a larger and more diverse sample set.

## Materials and Methods

### Clinical Sample Collection and Ethics

This research involved human participants and was performed in accordance with the relevant guidelines and regulations. Informed consent was obtained from all participants as required and was approved by Conjoint Health Research Ethics Board (CHREB) at the University of Calgary (REB18-0107, REB20-0402 and REB20-0444).

### RNA Extraction

All clinical and viral culture samples were extracted with the QIAGEN QIAmp Viral RNA Mini Kit following the kit protocol using centrifugation. A starting volume of 140 µL of UTM sample or culture fluid was used. RNA was eluted in 60 µL of AVE buffer. RNA extracts were aliquoted and frozen at −80 ºC after extraction so multiple ddPCR assays could be run on the same extracts while keeping the number of freeze-thaw cycles consistent so as to not compromise RNA quality.

### Viral Cultures

Vero Cells (ATCC#CCL-81) were maintained in Minimum Essential Media supplemented with 10% fetal bovine serum, 2 mM L-glutamine, 1 mM sodium pyruvate, 1x non-essential amino acids, and 1x antibiotics/antimycotics. Twenty-four hours before virus infection, Vero cells were seeded into 6-well plates at the density of 5E5 cells/well. For virus culture, Vero cells were infected at MOI = 0.01 in 500 µL inoculant/well. After one hour of absorption, another 2 mL of Minimum essential Media supplemented with 2 mM L-glutamine, 1 mM sodium pyruvate, 1x non-essential amino acids, and 1x antibiotics/antimycotics (SF-MEM) was added into each well. Virus culture were maintained at 37 ºC with 5% CO_2_. At designated time point, cells were scraped off from each well with media and centrifuged at 2000xg for 10 min. Supernatant (2 mL) were collected and the cell pellets and ∼0.5 mL media were collected as cell lysate. Cell lysates were frozen and thawed twice to release virus and RNAs from cells before assays. Vero cells were seeded at 2E5 cells/well in 24-well plates one day before the assay. One hundred microliter of inoculant from a serial 10-fold dilution were added in duplicated wells. After one hour of absorption, another 1 mL of SF-MEM with 1% carboxymethylcellulose was added into each well. Plates were maintained at 37 ºC with 5% CO_2_ for 3 days before fixation and staining. Cells were fixed/stained with a solution containing 0.13% Crystal violet, 5.26% ethanol, and 11.7% formaldehyde solution for at least one hour. Plaque counting were performed on an LED light box.

### Droplet Digital PCR (ddPCR)

ddPCR was performed on a Bio-Rad (Hercules, CA) system consisting of an AutoDG automated droplet generator, PX1 PCR plate sealer, C1000 Touch thermal cycler with 96–deep well reaction module, and a QX200 droplet reader. The details of each ddPCR assay used and evaluated in this study are listed in Table S2.

For the N1, N2, and RNase P assays, the Bio-Rad SARS-CoV-2 ddPCR kit was used according to the published instructions for use (19). All remaining ddPCR assays used the Bio-Rad One Step Advanced Kit for Probes as per the manufacturer protocol, modified only to use 9.9 μL of extracted RNA per reaction pre-AutoDG handling step. For the serial dilution of sample 1 and the quantification of samples 1, 6, 8, 9, and 10, the China CDC ORF1 ddPCR was run using RNA which went through an extra freeze-thaw step relative to the RNA used to quantify the other targets.

### Reverse Transcriptase PCR (RT-PCR)

RT-PCR was performed on a Bio-Rad CFX-96 real-time thermocycler. The same primers and probes used for ddPCR were used for RT-PCR with the exception of the US CDC N2 probe which was used entirely as a FAM probe for RT-PCR. For the Corman E gene RT-PCR, Many samples had less than 5 µL of RNA added to them (due to insufficient quantities remaining) and the RNA went through an extra freeze-thaw step compared to the other RT-PCR assays. Cycle threshold (Ct) values were calculated using the default method (single horizontal threshold for all reactions). Details of each assay are provided in Table S3.

### Data Analysis

Analysis of ddPCR data was performed using QX Manager 1.2 Standard Edition. Droplets were clustered manually in 2D amplitude mode. Graphs were made with MATLAB R2020b. Statistical analysis was performed in R Studio (R 4.1.2) and MATLAB R2020b. Permutational analysis of variance was performed with RVAideMemoire 0.9-80 (32).

## Data Availability

All data produced in the present study are available upon reasonable request to the authors

## Acknowledgments

We thank Claude Lachance and Ilya Grigoryev (Bio-Rad technical support) for help adapting RT-PCR assays for ddPCR.

## Author Contributions (CRediT Taxonomy)

Noah B. Toppings: Formal analysis, methodology, investigation, visualization, writing – original draft preparation, writing – review and editing.

Lisa K. Oberding: Formal analysis, methodology, investigation, writing – review and editing. Yi-Chan Lin: Formal analysis, investigation, writing – review and editing.

David Evans: Funding acquisition, conceptualization – review and editing.

Dylan R. Pillai: Supervision, funding acquisition, conceptualization, writing – review and editing.

## Supporting Information Legends

**Figure S1**. Standard curves of copies versus pfu for ddPCR targets using *in vitro* cultured SARS-CoV-2

**Table S1**. US CDC N2 RT-PCR Ct values of the eleven frozen clinical nasopharyngeal swabs used in this study.

**Figure S2**. Quantification of the Exact Diagnostics SARS-CoV-2 quantitative positive control by each ddPCR target.

**Figure S3**. Specific infectivity of SARS-CoV-2 cultured in Vero CCL81 cells.

**Table S2**. Details of ddPCR assays used in this study.

**Table S3**. Details of RT-PCR assays used in this study.

