## Supplementary Data for "The role of sub-genomic RNA in discordant results from RT-PCR tests for COVID-19"

**Supplementary Material:**

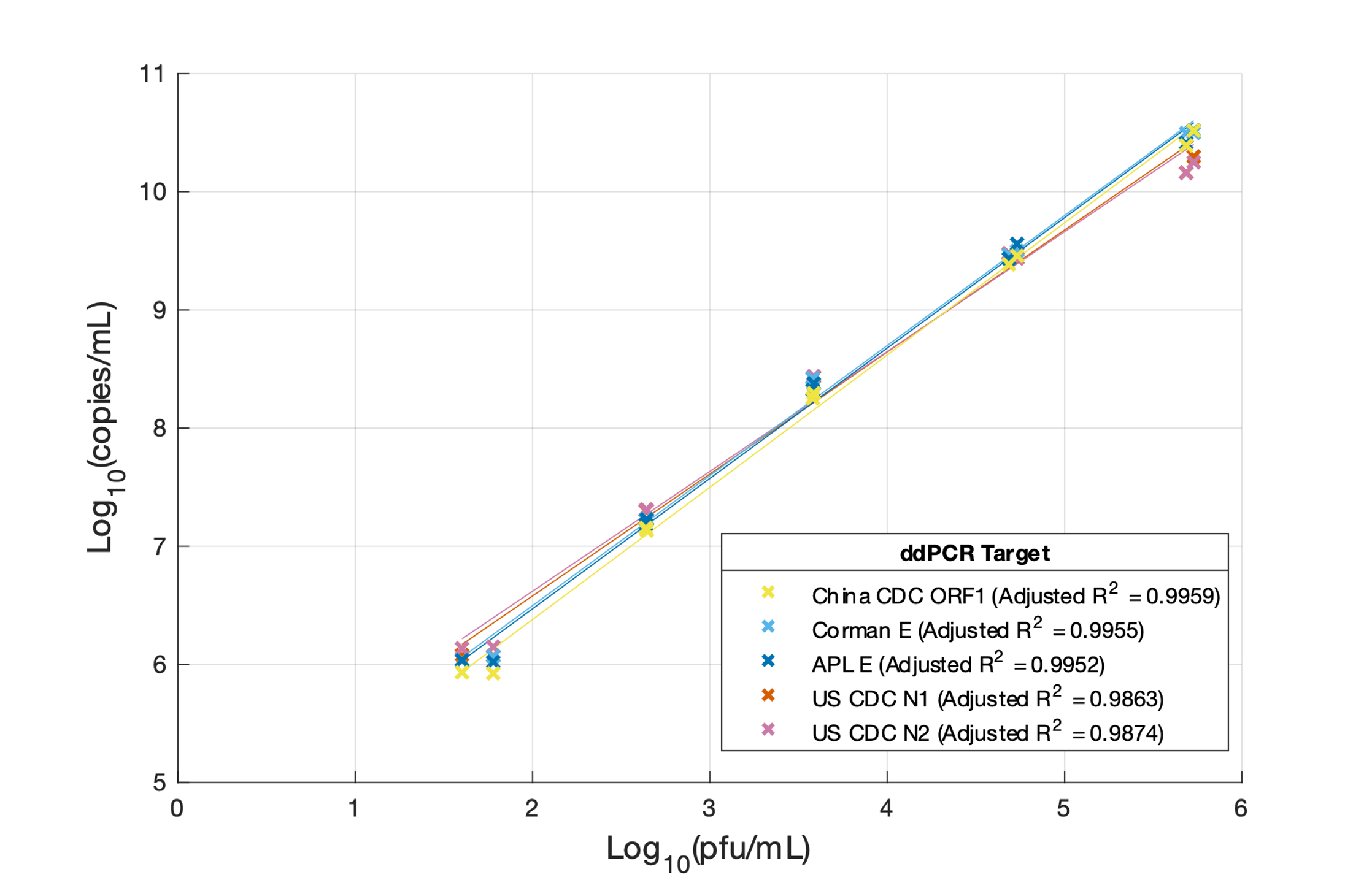

**Figure S1.** Standard curves of copies versus pfu for ddPCR targets using *in vitro* cultured SARS-CoV-2. Two cultures were grown and extracted at each of the five dilutions (n=10 per ddPCR target). RNA was extracted from culture supernatants.

**Table S1.** US CDC N2 RT-PCR Ct values of the eleven frozen clinical nasopharyngeal swabs used in this study.

| **Sample ID#** | **US CDC N2 gene RT-PCR Ct Value** |
| --- | --- |
| 1 | 19.04 |
| 2 | 20.76 |
| 3 | 21.89 |
| 4 | 23.82 |
| 5 | 25.8 |
| 7 | 27.11 |
| 6 | 27.16 |
| 9 | 29.46 |
| 11 | 30.21 |
| 8 | 30.28 |
| 10 | 31.95 |

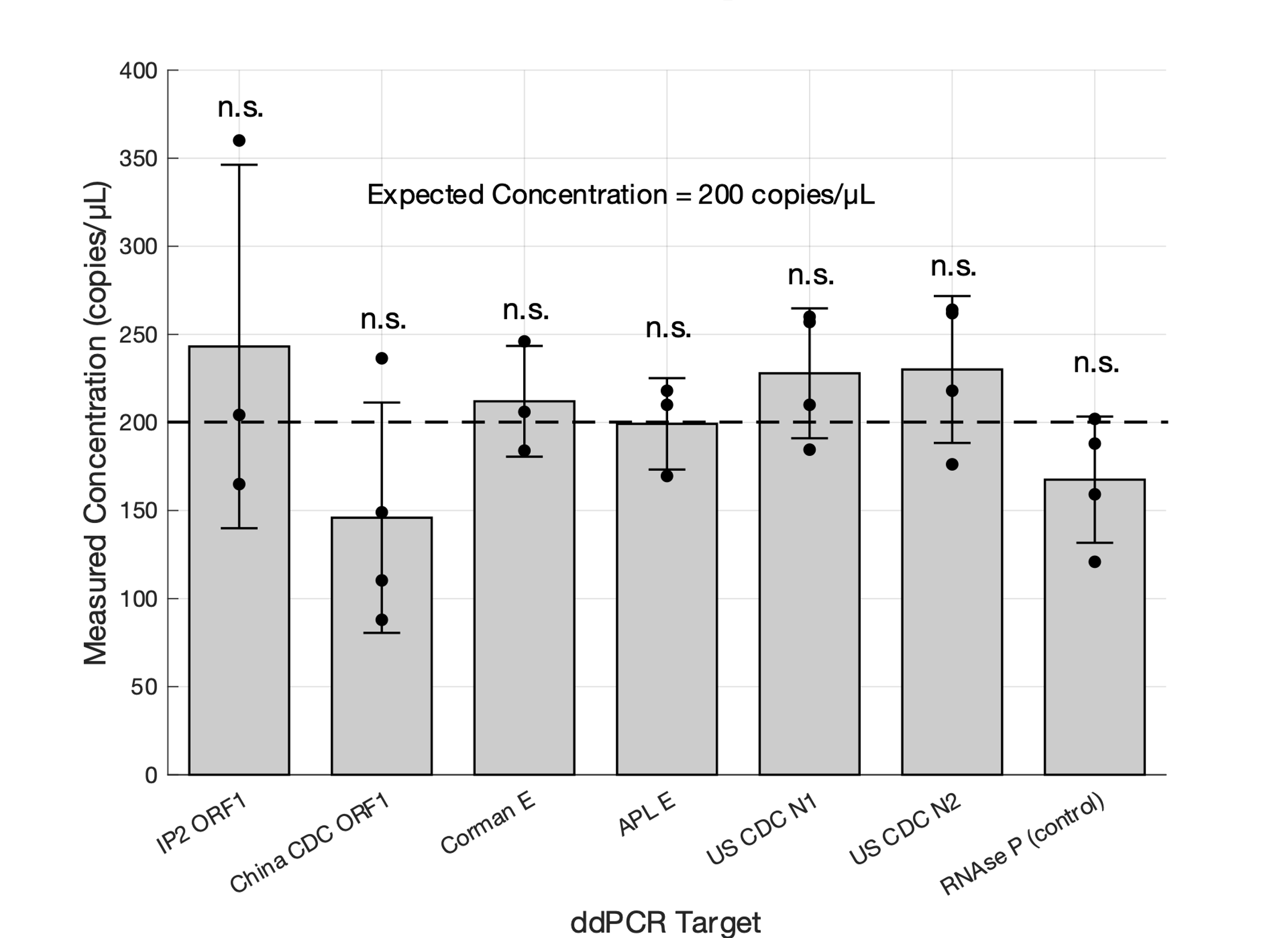

**Figure S2.** Quantification of the Exact Diagnostics SARS-CoV-2 quantitative positive control by each ddPCR target. N=4 for US CDC N1, US CDC N2, RNase P, and China CDC ORF1. N=3 for APL E, Corman E, and IP2 ORF1. All ddPCR assays quantitated the standard not significantly (n.s.) different than the expected concentration (single sample t-test, H_0_=200, p>0.05).

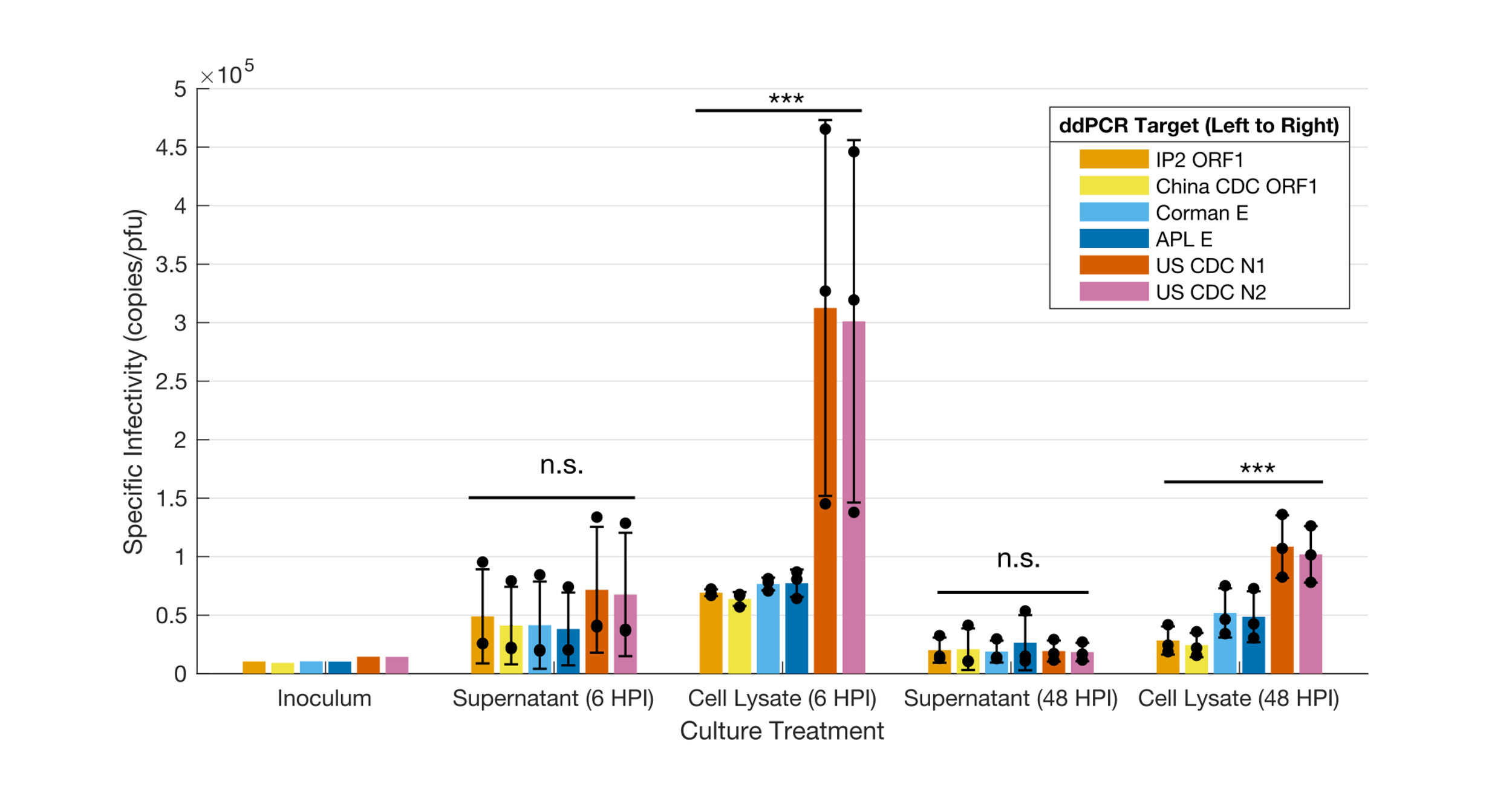

**Figure S3. Specific infectivity of SARS-CoV-2 cultured in Vero CCL81 cells.** Error bars show ± 1 standard deviation from the arithmetic mean of 3 replicates. An analysis of variance was conducted on each treatment (*** = p<0.001, n.s. = p>0.05). The supernatant (6 HPI) treatment was tested with the Kruskal-Wallis test. The cell lysate (6 HPI) was tested by permutation ANOVA test (1). The two 48 HPI treatments were tested with one-way ANOVA. The inoculum was not replicated (n=1). The mean counts from each treatment were used to make Figure 4B.

**Table S2.** Details of ddPCR assays used in this study. Primer and probe concentrations were used as per kit instructions. The thermocycling ramp rate used for all assays was 2 ºC/sec. †The US CDC N2 probe was a mixture of FAM and HEX to facilitate triplex detection by ddPCR along with N1 and RNase P (2).

| **ddPCR Target** | **Primer/Probe Set Author** | **ddPCR Thermocycling Profile Used** | **Primer/Probe** | **Sequence (5’🡪3’)** |
| --- | --- | --- | --- | --- |
| N1 | United States CDC (3) | Cycling Step \| Temperature (°C) \| Time \| Number of Cycles; Reverse Transcription \| 50 \| 60 min \| 1; Enzyme activation \| 95 \| 10 min \| 1; Denaturation \| 94 \| 30 sec \| 40; Annealing/Extension \| 55 \| 1 min \|; Enzyme Deactivation \| 98 \| 10 min \| 1; Hold and Droplet Stabilization \| 4 \| ∞ \| 1 From (2). | N1 Forward | GACCCCAAAATCAGCGAAAT |
|  |  |  | N1 Reverse | TCTGGTTACTGCCAGTTGAATCTG |
|  |  |  | N1 Probe | FAM – ACCCCGCATTACGTTTGGTGGACC – BHQ1 |
| N2 |  |  | N2 Forward | TTACAAACATTGGCCGCAAA |
|  |  |  | N2 Reverse | GCGCGACATTCCGAAGAA |
|  |  |  | N2 Probe† | FAM or HEX – ACAATTTGCCCCCAGCGCTTCAG – BHQ1 |
| RNase P |  |  | RNase P Forward | AGATTTGGACCTGCGAGCG |
|  |  |  | RNase P Reverse | GAGCGGCTGTCTCCACAAGT |
|  |  |  | RNase P Probe | FAM – TTCTGACCTGAAGGCTCTGCGCG – BHQ-1 |
| E gene | Corman *et al.* (4) | Cycling Step \| Temperature (°C) \| Time \| Number of Cycles; Reverse Transcription \| 50 \| 60 min \| 1; Enzyme activation \| 95 \| 10 min \| 1; Denaturation \| 95 \| 30 sec \| 45; Annealing/Extension \| 58 \| 1 min \|; Enzyme Deactivation \| 98 \| 10 min \| 1; Hold and Droplet Stabilization \| 4 \| ∞ \| 1 | Corman E Forward | ACAGGTACGTTAATAGTTAATAGCGT |
|  |  |  | Corman E Reverse | ATATTGCAGCAGTACGCACACA |
|  |  |  | Corman E Probe | FAM – ACACTAGCCATCCTTACTGCGCTTCG – BBQ |
| E gene | Alberta Precision Labs (APL) (5) | Cycling Step \| Temperature (°C) \| Time \| Number of Cycles; Reverse Transcription \| 50 \| 60 min \| 1; Enzyme activation \| 95 \| 10 min \| 1; Denaturation \| 95 \| 30 sec \| 41; Annealing/Extension \| 60 \| 1 min \|; Enzyme Deactivation \| 98 \| 10 min \| 1; Hold and Droplet Stabilization \| 4 \| ∞ \| 1 | APL E Forward | GAGACAGGTACGTTAATAGTTAATAGCG |
|  |  |  | APL E Reverse | CAATATTGCAGCAGTACGCACAC |
|  |  |  | APL E Probe | FAM – CTAGCCATCCTTACTGCG – MGB |
| ORF1ab (nsp10) | China CDC (6) | Cycling Step \| Temperature (°C) \| Time \| Number of Cycles; Reverse Transcription \| 50 \| 60 min \| 1; Enzyme activation \| 95 \| 10 min \| 1; Denaturation \| 95 \| 30 sec \| 45; Annealing/Extension \| 60 \| 1 min \|; Enzyme Deactivation \| 98 \| 10 min \| 1; Hold and Droplet Stabilization \| 4 \| ∞ \| 1 | China CDC ORF1 Forward | CCCTGTGGGTTTTACACTTAA |
|  |  |  | China CDC ORF1 Reverse | ACGATTGTGCATCAGCTGA |
|  |  |  | China CDC ORF1 Probe | FAM – CCGTCTGCGGTATGTGGAAAGGTTATGG – BHQ1 |
| IP2 (ORF1, also known as RdRP) | Institute Pasteur, Paris, France (7) | Cycling Step \| Temperature (°C) \| Time \| Number of Cycles; Reverse Transcription \| 50 \| 60 min \| 1; Enzyme activation \| 95 \| 10 min \| 1; Denaturation \| 94 \| 30 sec \| 45; Annealing/Extension \| 52.2 \| 1 min \|; Enzyme Deactivation \| 98 \| 10 min \| 1; Hold and Droplet Stabilization \| 4 \| ∞ \| 1 | IP2 ORF 1 Forward | ATGAGCTTAGTCCTGTTG |
|  |  |  | IP2 ORF 1 Reverse | CTCCCTTTGTTGTGTTGT |
|  |  |  | IP2 ORF1 IP2 Probe | HEX – AGATGTCTTGTGCTGCCGGTA – BHQ-1 |

**Table S3.** Details of RT-PCR assays used in this study. The same primers and probes used for ddPCR were used for RT-PCR with the exception of the US CDC N2 probe which was used entirely as a FAM probe for RT-PCR. All RT-PCR reactions were ran on a Bio-Rad CFX-96 real-time thermocycler. †Many samples had less than 5 µL of RNA added to them (due to insufficient quantities remaining) and the RNA went through an extra freeze-thaw step compared to the other RT-PCR assays.

| **RT-PCR Target** | **Reaction Mix Setup** | **Thermocycling Profile** |
| --- | --- | --- |
| US CDC N1 (8) | 8.5 µL nuclease free water, 1.5 µL combined primer/probe mix (IDT), 5 µL TaqPath 1-Step RT-qPCR Master Mix (4X), 5 µL RNA sample. | Temperature (°C) \| Time \| Number of Cycles; 25 ºC \| 2 min \| 1; 50 ºC \| 15 min \|; 95 ºC \| 2 min \|; 95 ºC \| 3 sec \| 45; 55 ºC \| 30 sec \| |
| US CDC N2 (8) |  |  |
| Corman E (9) | 11.25 µL nuclease free water, 1 µL forward primer (10 µM), 1 µL reverse primer (10 µM), 0.5 µL probe (10 µM), 6.25 µL TaqPath 1-Step RT-qPCR Master Mix (4X), 5 µL RNA sample†. | Temperature (°C) \| Time \| Number of Cycles; 55 ºC \| 10 min \| 1; 94 ºC \| 3 min \|; 94 ºC \| 15 sec \| 45; 58 ºC \| 30 sec \| |
| APL E (5) | 1.5 µL nuclease free water, forward primer 0.4 µL (20 µM stock), reverse primer 0.4 µL (20 µM stock), probe 0.2 µL (10 µM stock), Taqman Fast Virus One-Step RT-PCR Master Mix (4X), 5 µL RNA sample. | Temperature (°C) \| Time \| Number of Cycles; 50 \| 5 min \| 1; 95 \| 20 sec \|; 95 \| 5 sec \| 45; 60 \| 30 sec \| |
| China CDC ORF1 (6) | 7.5 µL nuclease free water, 1 µL forward primer (10 µM stock), 1 µL reverse primer (10 µM stock), 0.5 µL probe (10 µM stock), 5 µL Taqman Fast Virus One-Step RT-PCR Master Mix (4X), 5 µL RNA sample. |  |
| IP2 ORF1 (7) | 11.25 µL of nuclease free water, 1 µL forward primer (10 µM stock), 1 µL reverse primer (10 µM stock), 0.5 µL probe (10 µM stock), 6.25 µL Taqman Fast Virus One-Step RT-PCR Master Mix (4X), 5 µL RNA sample. | Temperature (°C) \| Time \| Number of Cycles; 55 ºC \| 20 min \| 1; 95 ºC \| 3 min \|; 95 ºC \| 15 sec \| 50; 58 ºC \| 30 sec \|; 40 ºC \| 30 sec \| 1 |

6.	World Health Organization (WHO). WHO In House Assays. 2021.

7.	Institute_Pasteur. Protocol: Real-time RT-PCR assays for the detection of SARS-CoV-2. 2020.

8.	Centers for Disease Control and Prevention. CDC 2019-Novel Coronavirus (2019-nCoV) Real-Time RT-PCR Diagnostic Panel For Emergency Use Only Instructions for Use Revision 01 [Internet]. 2020 [cited 2020 Nov 4]. Available from: http://www.mlpla.mil.cn/dzfw/yhjy/xgzl/202002/P020200210749561366112.pdf

9.	Corman VM, Landt O, Kaiser M, Molenkamp R, Meijer A, Chu DK, et al. Detection of 2019 novel coronavirus (2019-nCoV) by real-time RT-PCR. Eurosurveillance. 2020 Jan 23;25(3):2000045.
